# Comparison of IgG and neutralizing antibody responses after one or two doses of COVID-19 mRNA vaccine in previously infected and uninfected persons

**DOI:** 10.1101/2021.03.04.21252913

**Authors:** Alexis R. Demonbreun, Amelia Sancilio, Matt E. Velez, Daniel T. Ryan, Rana Saber, Lauren A. Vaught, Nina L. Reiser, Ryan R. Hsieh, Richard T. D’Aquila, Brian Mustanski, Elizabeth M. McNally, Thomas W. McDade

## Abstract

**Objective:** To compare anti-SARS-CoV-2 spike receptor binding domain (RBD) IgG antibody concentrations and antibody-mediated neutralization of spike-ACE2 receptor binding *in vitro* following vaccination of non-hospitalized participants by sero-status and acute virus diagnosis history.

**Methods:** Participants were studied before and after mRNA vaccination in a community-based, home-collected, longitudinal serosurvey; none reported hospitalization for COVID-19. Prior to vaccination, some reported prior positive acute viral diagnostic testing and were seropositive (COVID-19+). Participants who did not report acute viral diagnostic testing were categorized as seropositive or seronegative based on anti-spike RBD IgG test results. Primary measures were anti-spike RBD IgG concentration and percent antibody-mediated neutralization of spike protein-ACE2 interaction prior to vaccination, and after one or two doses of vaccine.

**Results:** Of 290 unique vaccine recipients, 42 reported a prior COVID-19 diagnosis and were seropositive (COVID-19+). Of the 248 with no history of acute viral diagnostic testing, 105 were seropositive and 143 seronegative before vaccination. The median age was 38yrs (range 21-83) with 65% female and 35% male; 40% were non-white. Responses were evaluated after one (n=140) or two (n=170) doses of BNT162b2/Pfizer or mRNA-1273/Moderna vaccine. After one dose, median post-vaccine IgG concentration and percent neutralization were each significantly higher among the COVID-19+ group (median 47.7 µg/ml, IgG; >99.9% neutralization) compared to the seropositives (3.4 µg /ml IgG; 62.8% neutralization) and seronegatives (2.2 µg /ml IgG; 39.5% neutralization). The latter two groups reached >95% neutralization after the second vaccine dose.

**Conclusions:** A prior outpatient COVID-19 diagnosis was associated with strong anti-spike RBD IgG and *in vitro* neutralizing responses after one vaccine dose. Persons seropositive for anti-spike RBD IgG in the absence of acute viral diagnostic testing, and those who were seronegative, required two doses to achieve equivalently high levels of IgG and neutralization activity. One mRNA vaccine dose is not sufficient to generate *in vitro* evidence of strong protection against COVID-19 among most persons previously infected with SARS-CoV-2, nor among seronegative persons.

## INTRODUCTION

In December 2020, the FDA authorized the emergency use of two SARS-CoV-2 spike mRNA vaccines that utilize a 2-dose vaccine schedule administered several weeks apart: BNT162b2/Pfizer and mRNA-1273/Moderna ^1,2^. While phase 3 trials for both vaccines reported high efficacy in preventing symptomatic SARS-CoV-2 infections after administration of the second dose, recent reports have suggested that a single dose is sufficient to boost immunity to full protection among previously infected individuals ^3-5^. Since anti-SARS-CoV-2 seropositivity significantly exceeds documented COVID-19 cases in the community ^6,7^, vaccine supply could be immediately increased if second doses were not needed for a large percentage of the population.

In this study, we compared antibody response to vaccination across three groups: individuals who previously recovered from a documented outpatient COVID-19 infection and were anti-SARS-CoV-2 spike receptor binding domain (RBD) seropositive, individuals who were seropositive for anti-RBD IgG but who had no acute virus diagnostic test for COVID-19, and individuals who both lacked acute virus diagnostic testing and were seronegative prior to vaccination. The magnitude of anti-RBD IgG response, and percentage neutralization of spike-ACE2 receptor binding—an *in vitro* surrogate for protection following vaccination—were quantified among participants in a community-based seroprevalence study ^6,7^. We document robust responses to the first vaccine dose in the recovered COVID-19 group, but more heterogeneous and modest responses among those not reporting acute virus diagnostic testing who were either seropositive or seronegative. Results suggest caution in assuming immunological priming and enhanced responses to a first mRNA vaccine dose based on the presence of anti-SARS-CoV-2 antibodies alone.

## METHODS

### Study approvals

Research activities were implemented under conditions of informed consent with protocols approved by the institutional review board at Northwestern University (#STU00212457, and #STU00212472). All samples and data were de-identified.

### Participants and study design

Approximately 8000 pre-vaccinated participants were recruited to Screening for Coronavirus Antibodies in Neighborhoods (SCAN) from across the Chicagoland area from April to December 2020. Eligible participants consented online and completed a questionnaire regarding COVID-19 status and symptoms. Participants received and returned materials for finger-stick dried blood spot (DBS) collection through the mail or on-site pick up. From December 2020 through January 2021, participants were contacted at the email address they had provided and queried regarding COVID-19 vaccination status. Participants were resampled with additional DBS self-collected at home through February 2021 if they reported receiving one or two doses of either mRNA vaccine. We excluded DBS collected earlier than 9 days after dose 1 or earlier than day 5 after dose 2. Participants were categorized as recovered COVID-19 if they reported testing positive for SARS-CoV-2 on a clinical molecular diagnostic test for acute infection any time prior to vaccination. Participants who lacked self-report of a positive SARS-CoV-2 clinical diagnostic test result were categorized as seropositive or seronegative based on the presence of anti-RBD IgG antibodies in DBS samples collected between April and December 2020.

### Serological assay

The enzyme linked immunosorbent assay (ELISA) protocol to measure anti-RBD IgG levels was previously described ^8,9^. DBS samples were run in duplicate and reported as the average. Results were normalized to the CR3022 antibody with known affinity to RBD ^10^. DBS sample anti-RBD IgG concentration (µg/ml) was calculated from the four parameter logistic regression of the CR3022 calibration curve. A value >0.39µg/ml CR3022 was considered positive. Samples with high IgG required 1:16 dilution prior to ELISA.

### Surrogate virus neutralizing assay

The competitive immunoassay to quantify neutralizing activity (% neutralization) of spike-ACE2 interaction was previously described ^11^. DBS samples were incubated with SARS-CoV-2 spike protein and ACE2 conjugated with an electrochemiluminescent label. Neutralizing antibodies, if present, inhibited binding between ACE2 and spike protein, and the Meso Scale Diagnostics QuickPlex SQ 120 Imager was used to read mean fluorescence intensity (MFI). DBS samples were run in duplicate and reported as the average. Percent neutralization was calculated as follows: % neutralization = 100 × 1-(sample MFI/negative control MFI).

### Statistical Analysis

Descriptive statistics and two-way Mann-Whitney tests were used to evaluate differences in antibody concentration and % neutralization across groups. P<0.05 was set as the criterion for statistical significance.

## RESULTS

The study groups were comparable in terms of age, gender, race/ethnicity, and duration of days between vaccination and blood sampling (**Table 1**). The convalescent COVID-19+ group reported a median of five (interquartile range (IQR): 2-7) symptoms of COVID-19 infection and had a median interval of 138 days between their positive acute SARS-CoV-2 infection diagnostic test and vaccination. The seropositive group, with no prior acute virus testing, reported a median of one symptom (IQR: 0-3), (z=4.51, p<0.001). Prior to vaccination, median anti-RBD IgG concentration (z=3.72, p<0.001) and % neutralization (z=4.62, p<0.001) were significantly lower for seropositive participants in comparison with the recovered COVID-19-positive group.

**Table 1.**
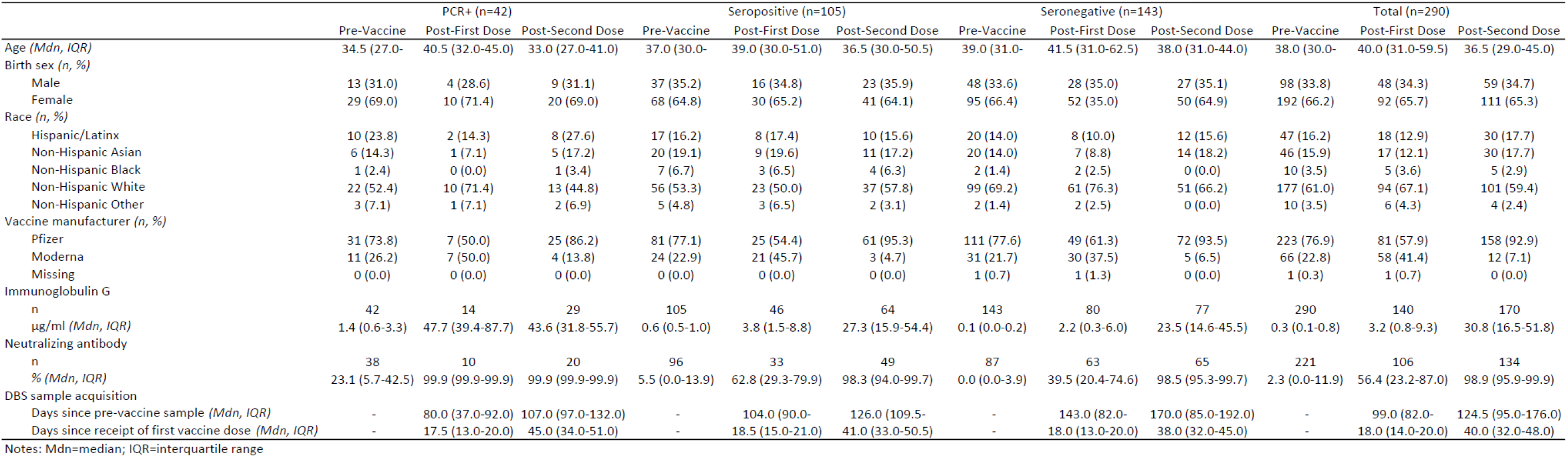
Sample characteristics by pre-vaccine serology and timing of dried blood spot sample collection

Following the first vaccine dose, there were large increases in anti-RBD IgG and neutralization activity for convalescent COVID-19+ cases, and modest increases for the seropositive group (**Figure 1**). In the COVID-19+ group, median IgG increased from baseline by a factor of 34.8, and all samples exceeded 95% neutralization. By contrast, in the seropositive group, anti-RBD IgG increased by a factor of 5.3 and was significantly lower than the recovered COVID-19 positive group (z=5.21, p<0.001). Median % neutralization was also significantly lower at 62.8% (z=4.28, p<0.001), with only 5 of 33 samples reaching >95% neutralization. The seropositive and seronegative groups did not differ in % neutralization following the first vaccine dose (z=1.72, p>0.05) and there was a small but significant difference in anti-RBD IgG between them (z=2.11, p<0.05).

**Figure 1:**
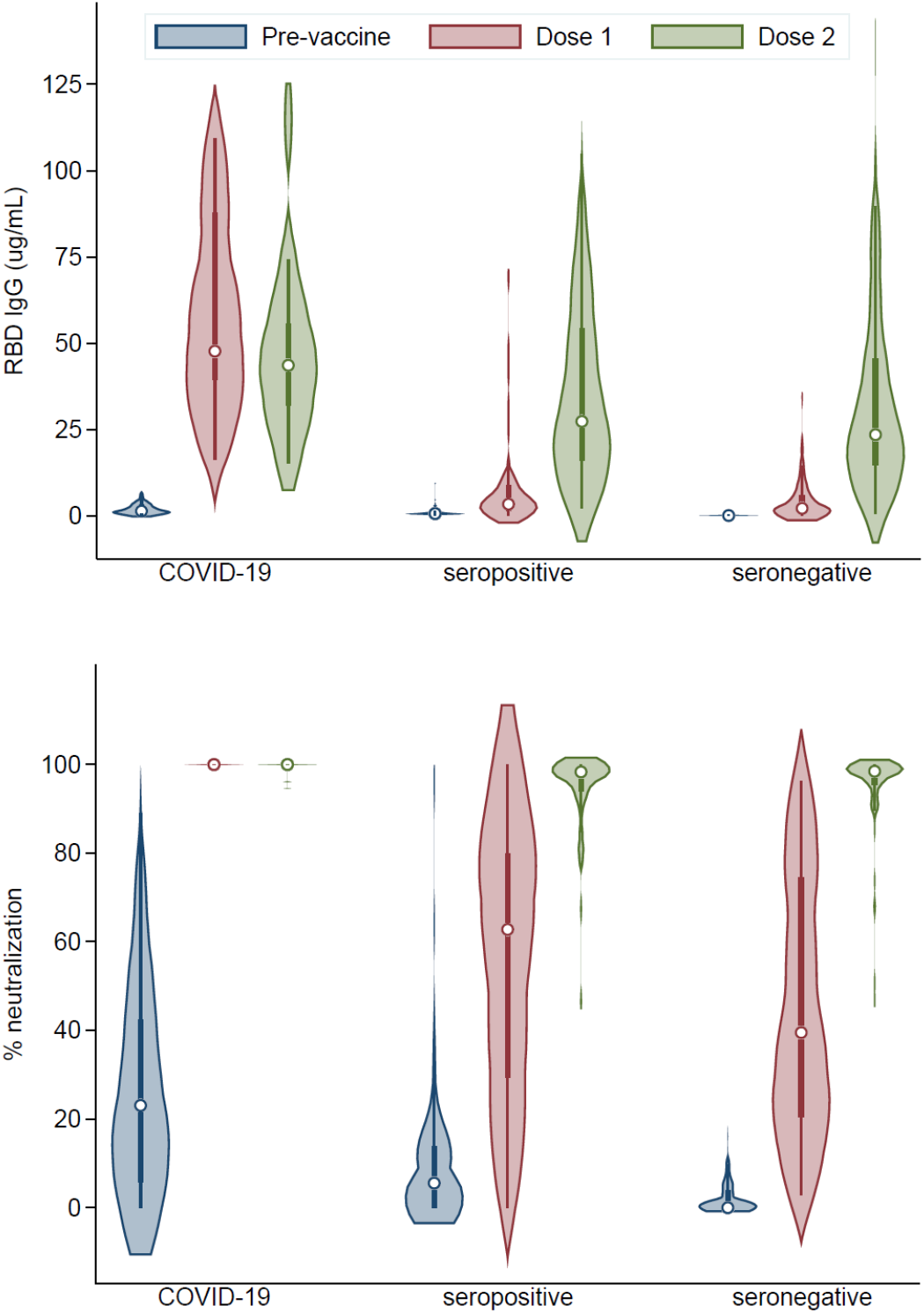
Pattern of anti-RBD IgG and neutralization antibody response to vaccination across study groups. Violin plots show median level and interquartile range, with kernel density, before vaccination, after dose 1, and after dose 2, for three groups: convalescent COVID-19, seropositive for SARS-CoV-2, and seronegative. The COVID-19 group had significantly higher pre-vaccine median IgG (1.37 vs. 0.64 ug/mL, p<0.001) and median % neutralization (23.1 vs. 5.5, p<0.001) in comparison with the seropositive group. The COVID-19 group had significantly higher post-dose 1 median IgG (47.71 vs. 3.38 ug/mL, p<0.001) and median % neutralization (99.9 vs. 62.8, p<0.001) in comparison with the seropositive group. The seropositive group had significantly higher post-dose 1 median IgG (3.37 vs. 2.16 ug/mL, p<0.05) than the seronegative group, but the groups did not differ in median % neutralization (62.8 vs. 39.5, p=0.09). The COVID-19 group had significantly higher post-dose 2 median IgG (43.60 vs. 27.34 ug/mL, p<0.05) and median % neutralization (99.9 vs. 98.3, p<0.001) in comparison with the seropositive group. The seropositive and seronegative groups did not differ significantly in post-dose 2 median IgG (27.34 vs. 23.50, p=0.43) or median % neutralization (98.3 vs. 98.5, p=0.72).

Following the second vaccine dose, the convalescent COVID-19+ group had anti-RBD IgG and % neutralization that were comparable to the high levels recorded after the first dose. In the seropositive group, anti-RBD IgG was 8.1 times higher than after the first dose, but still significantly lower than the level observed in the COVID-19+ group (z=2.37, p<0.05). Percent neutralization was 1.6 times higher than after the first dose, but median neutralization remained significantly lower than the COVID-19+ group (z=4.73, p<0.001). Fifteen of 50 (30.0%) seropositive individuals did not exceed 95% neutralization after the second dose within our sampling timeframe. The seropositive and seronegative groups did not differ significantly in anti-RBD IgG concentration (z=0.79, p=0.43) or % neutralization (z=-0.36, p=0.72) after a second dose.

Anti-RBD IgG concentration positively correlated with % neutralization for all groups after one dose of vaccine, with a wide range of values for the seropositive and seronegative groups in comparison with the more consistently high levels among the recovered COVID-19+ group (**Figure 2**).

**Figure 2:**
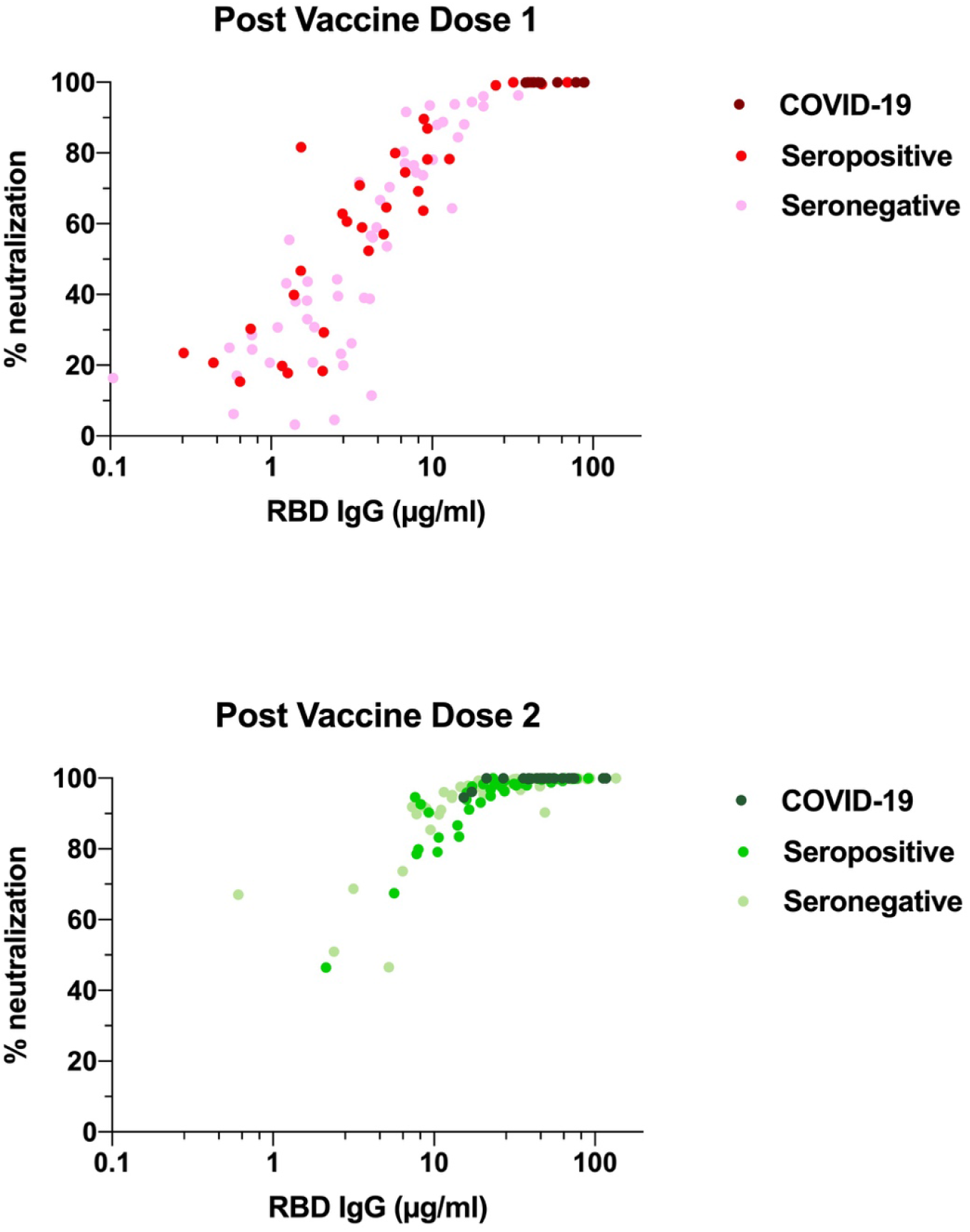
Percentage antibody-mediated neutralization positively correlates with anti-RBD IgG concentration. Dried blood spots samples were evaluated for both anti-RBD IgG levels and % antibody-mediated neutralization of spike protein-ACE2 interaction. After one dose of COVID-19 vaccine, a wide range of IgG concentrations and % neutralization were observed in seropositive and seronegative samples, while all recovered COVID-19 participants reached >95% neutralization (r = 0.73 COVID-19+ n=9; r= 0.90 seropositive n= 32; r= 0.86 seronegative n=61). Anti-RBD IgG concentration and % neutralization increased dramatically after two doses of vaccine in the majority of seropositive and seronegative samples (r = 0.83 COVID-19+ n=20; r= 0.90 seropositive n= 49; r= 0.89 seronegative n=66).

## DISCUSSION

Following the initial phase of vaccine deployment it has been suggested that two doses of currently available mRNA vaccines are not necessary for individuals previously diagnosed with COVID-19, and for those who test seropositive for SARS-CoV-2 ^3,4^. If a single dose is sufficient to boost protection among these groups then vaccine supplies can be extended to vaccinate more of the population in a shorter amount of time. We document strong antibody responses to the first vaccine dose among individuals with confirmed cases of COVID-19, consistent with recent reports ^3^. We document a pattern of mild and heterogeneous responses to the first dose among individuals previously unexposed to SARS-CoV-2, with more robust responses following the second dose, consistent with clinical trials data ^2^. Importantly, responses in the seropositive group suggest that immunity following the first vaccine dose is significantly lower than the convalescent COVID-19 group. And like the seronegative group, two doses are required for the seropositive group to attain a level of protection that is comparable to the COVID-19 group.

The majority of cases of COVID-19 are asymptomatic, or minimally symptomatic and require only home-based treatment ^7,12^. These milder cases are known to produce lower levels of antibodies. Studies of vaccine effectiveness that focus on convalescent patients who were hospitalized for more serious COVID-19, or samples enriched with people at higher risk of exposure and potentially repeated exposures (e.g., health care workers), may lead to overestimates of the level of priming immunity that do not generalize to the entire population of seropositive individuals with less intense or shorter exposure to SARS-CoV-2 antigens during natural infection. Results from this community-based study, with clinically confirmed symptomatic as well as asymptomatic/mild cases of infection, suggest that seropositivity alone does not indicate sufficient memory to generate high levels of protection following a single dose of the BNT162b2/Pfizer and mRNA-1273 vaccines.

## Data Availability

Data available upon reasonable request from corresponding author to protect participant confidentiality.

